# Development and validation of a length- and habitus-based method of ideal and lean body weight estimation for critically ill adults requiring urgent weight-based medical intervention

**DOI:** 10.1101/2021.10.04.21264460

**Authors:** Mike Wells, Lara Goldstein, Giles Cattermole

**Author notes:** Corresponding author: Prof M Wells, 11200 SW 8th Street, Miami, FL 33199.

## Abstract

**Objective:** Accurate drug dosing in obese patients requires an estimation of ideal body weight (IBW) or lean body weight (LBW) for dosing hydrophilic medications. Erroneous weight estimates during the management of critically ill adults may contribute to poor outcomes. Existing methods of IBW and LBW estimation or measurement are very difficult to use during emergency care. A new point-of-care model is needed to provide rapid estimates of IBW and LBW for this purpose.

**Methods:** A model was derived based on the PAWPER XL-MAC tape, a paediatric weight estimation system, which uses recumbent length and mid-arm circumference to estimate IBW and LBW. The model was used to generate weight estimations in a derivation sample (n=33155) and a validation sample (n=5926) from National Health and Nutrition Examination Survey (NHANES) datasets. The outcome measure was to achieve >95% of IBW and LBW estimations within 20% of recognised reference standards (P20>95%) and >70% of estimations within 10% of these standards (P10>70%).

**Main Results:** The new model achieved a P20 of 100% and a P10 of 99.9% for IBW and a P20 of 98.3% and a P10 of 78.3% for LBW. This accuracy was maintained in both sexes, all ages, all ethnic groups, all lengths and in all habitus-types, except for the morbidly obese female subgroup.

**Conclusions:** The modified PAWPER XL-MAC model proved to be an accurate method of IBW and LBW estimation. It could, therefore, have an important role in facilitating emergency drug dose calculations in acutely or critically ill obese adult patients.

**Article summary:** *Why is this topic important?:* Errors in weight estimation, or the use of an inappropriate weight scalar, can translate into medication errors and potential patient harm. The ability to easily estimate TBW, IBW and LBW would enhance patient safety and make correct drug dosing more accessible during the provision of emergency and critical care.

*What does this study attempt to show?:* This study describes the derivation and validation of a point of care model to provide estimates of IBW and LBW for the purposes of weight based dosing. The model is an adult version of a well-studied paediatric weight estimation system.

*What are the key findings?:* The model was able to provide accurate estimations of IBW and LBW in a wide range of patient-types. It was the first study to have evaluated such a system. It established that this system would be useful to test in clinical trials.

*How is patient care impacted?:* The PAWPER XL-MAC system could provide quick and easy estimations of IBW and LBW which can be used in obese patients for appropriate drug dose calculations. This would improve the accuracy of dosing with a potential reduction in medication errors and patient harm.

## INTRODUCTION

Critically ill or injured obese patients are at high risk when it comes to the dose calculations involving potentially life-saving treatments [1]. Paradoxically, these patients are at risk of underdosing as well as overdosing of medications, if the various drugs administered are not scaled to their appropriate weight descriptor [2]. Furthermore, abnormalities in cardiac and respiratory function resulting from obesity, as well as the effects of critically altered physiology on pharmacokinetics, may all narrow the therapeutic window of high-risk drugs, increasing the jeopardy of dosing errors [3]. It has been reasonably well established that (in general) lipophilic drugs should be dosed to total body weight (TBW) in obese patients to prevent underdosing, but hydrophilic drugs should be dosed to ideal body weight (IBW) or lean body weight (LBW) to avoid overdosing [4]. The use of the correct scalar is, therefore, important for patient safety.

The real-world problem is that it is very hard to estimate IBW and LBW with existing methods as they require virtually impossible-to-remember, complex formulas [5]. LBW also cannot be directly measured in critically or acutely ill patients as there is no validated easy-to-use point-of-care method for this purpose. This raises the question about the potential harm that is incurred by obese patients during emergency and critical care when weight-based drug therapy is not individualised appropriately. While TBW can sometimes be measured by weighing critically ill patients, it may be difficult to accomplish timeously, and – somewhat surprisingly – has not been well validated [6, 7]. Therefore, a method to estimate TBW and IBW or LBW is required to facilitate weight-based medication dose calculations.

Since the prevalence of obesity exceeds 30% in many countries worldwide (including the United States) it is inevitable that Emergency Physicians, Anaesthetists, and Intensivists will be faced with managing critically ill obese patients on every shift [8]. It is, therefore, important to be able to determine drug doses both accutely and easily in these patients. The ability to estimate the correct dose according to the patient’s body habitus using the appropriate weight scalar might make the difference between treatment failure (dose too small), efficacious treatment (dose just right), and overdose with potentially harmful side effects (dose too large) [9]. However, using different weight descriptors for different drugs during the emergency care of obese patients may increase the complexity of the process of determining drug doses under already cognitively stressful circumstances. For this reason, such a weight-estimation system must be simple and easy to use [10].

There is recent evidence that the best of the dual length- and habitus-based paediatric weight estimation systems can be successfully used in adults to estimate TBW [11-13]. One of these methods is the PAWPER XL-MAC tape which makes use of recumbent body length and mid-arm circumference (MAC) to generate estimates of weight, which can be read directly off the tape. It outperformed all other methods of TBW estimation, except for patient self-estimations [11]. The PAWPER XL-MAC method has also been shown to be able to predict IBW accurately in children [14, 15]. In adults, since IBW is closely related to height and LBW is closely related to height and TBW, it is possible that the PAWPER XL-MAC system could be adapted to predict IBW and LBW in addition to TBW [16].

We hypothesized that the PAWPER XL-MAC method could be adapted for use in adults, to produce accurate, easy-to-use estimates of IBW and LBW. The aim of this study was, therefore, to develop and validate an adult version of the PAWPER XL-MAC tape, using recumbent length and MAC to predict IBW and LBW.

## METHODS

The model that was developed was based on the PAWPER XL-MAC tape paediatric weight estimation system (see Figure 1) [17]. The process used for developing, calibrating, and validating the new model is shown in Figure 2. The devised system has already been validated for estimations of TBW (Wells M, Goldstein LN, Cattermole G. Development and validation of a length- and habitus-based method of total body weight estimation for critically ill adults requiring urgent weight-based medical intervention. Submitted for publication).

**Fig 1.**
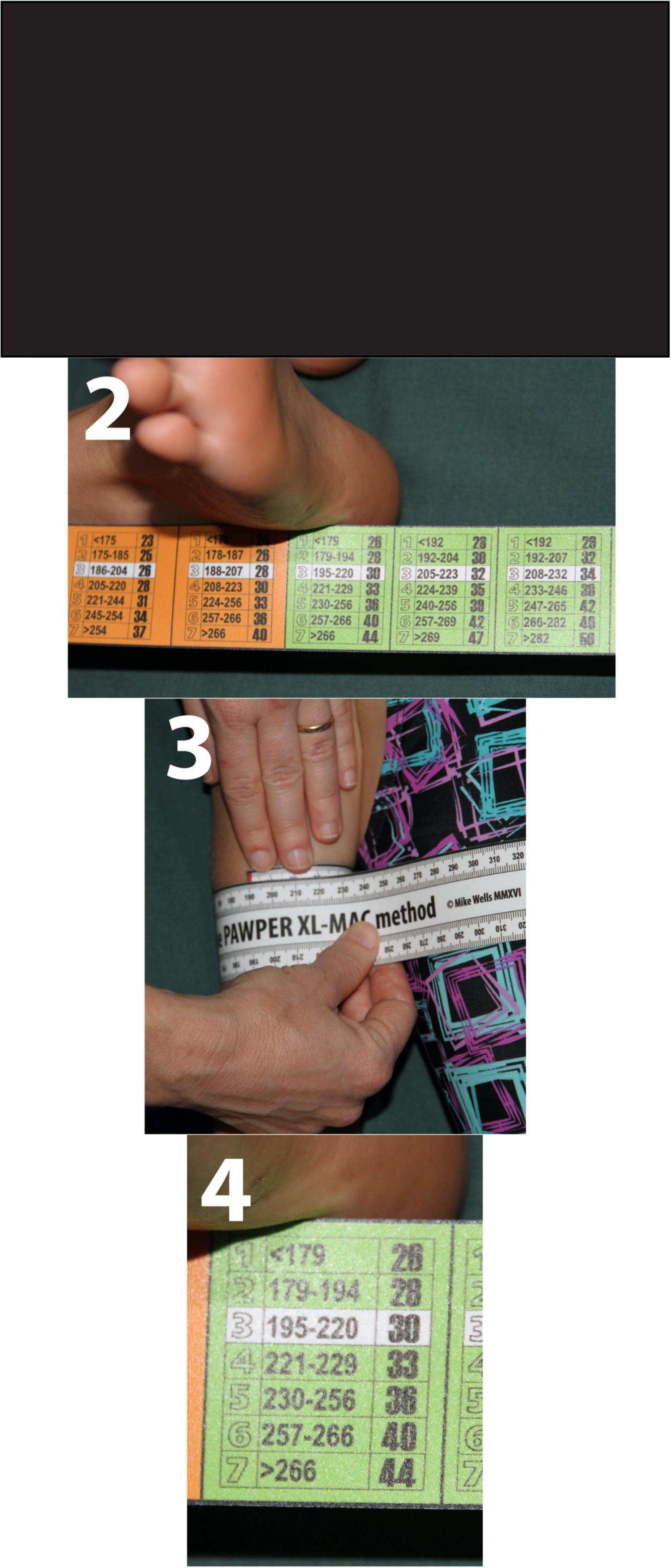
How to use the PAWPER XL-MAC tape. The method of using the tape is the same for children and adults. Panel 1 – the tape is stretched from head to heel using a one-person or, more ideally, a two-person technique. Panel 2 – the segment at which the patient’s heel crosses the line is noted. Panel 3 – mid-arm circumference is measured using the scale provided on the tape itself. Panel 4 – this measurement can be applied in the length segment determined in Step 2. The weight can then be read directly off the tape. This tape only shows total body weight, but the adult version of the tape will have total body weight, ideal body weight and lean body weight data.

**Fig 2.**
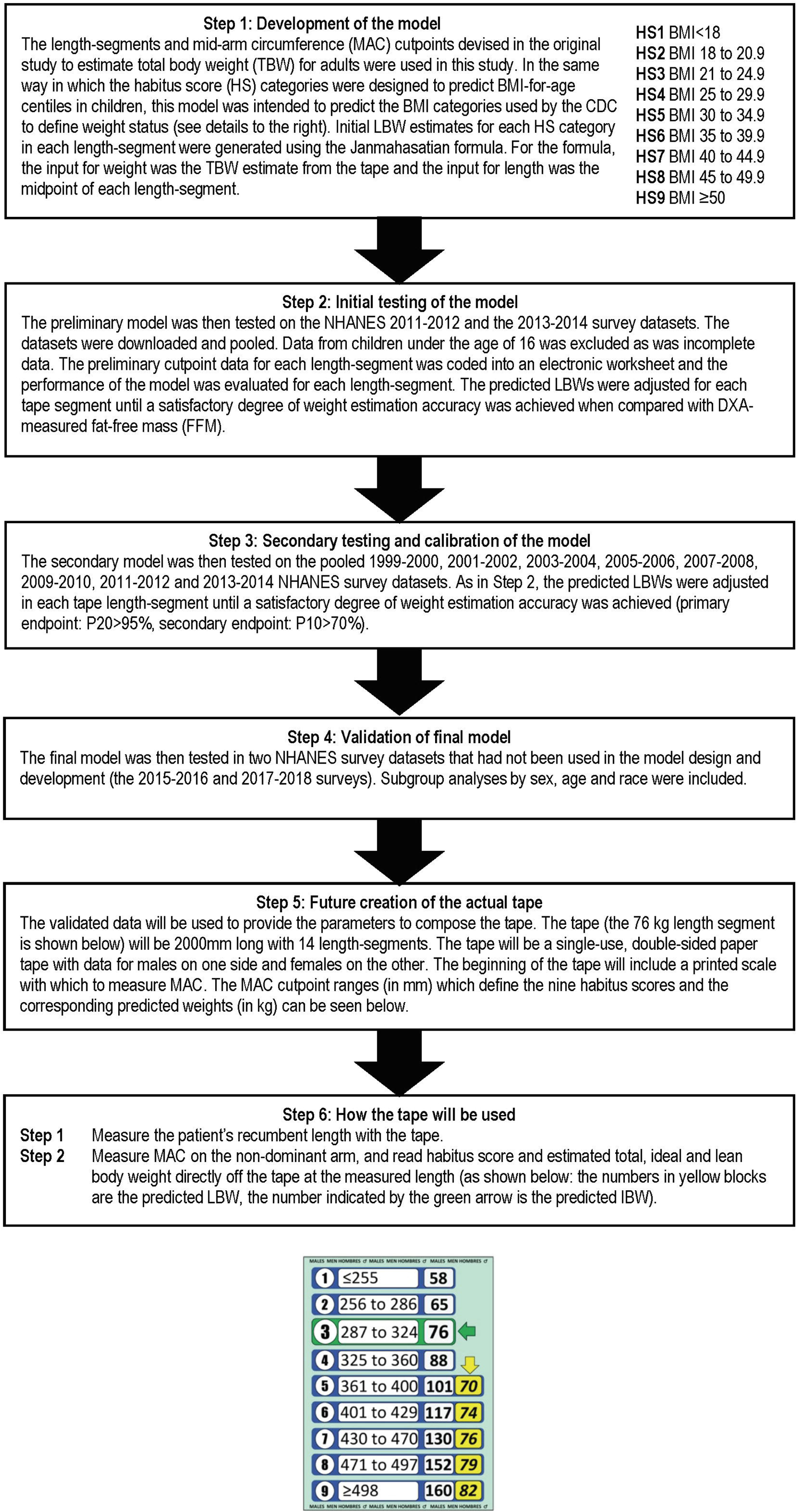
The process used in the development, calibration, and validation of the new PAWPER XL-MAC method for estimating ideal body weight and lean body weight in adults. Although lean body weight (LBW) and fat-free mass (FFM) are often used interchangeably, there are subtle differences. These differences are not important for dose calculations. In this paper LBW is used when referring to a predicted weight, while FFM is used when referring to a DXA-measured weight.

### Method development

The existing length-segments, MAC cutpoint values and habitus score (HS) categories of the original paediatric version of the PAWPER XL-MAC tape were used for the provisional model. New length-segments were added to extend the length of the tape from 1800 mm to 2000 mm. The MAC cutpoints to define the HS categories were extrapolated from data in the proximal segments. Initial TBW estimates for each HS category were extrapolated from weights obtained from the Centers for Disease Control and Prevention (CDC) weight-for-height charts in each length-segment (HS1 from the 15^th^ centile, HS2 from the 25^th^ centile, HS3 from the 50^th^ centile, HS4 from the 85^th^ centile, HS5 from the 95^th^ centile, HS6 from the 97.5^th^ centile and HS7 from the 99^th^ centile). IBW estimates for each segment were defined to be predicted by the HS3 weight (“normal” weight) in that segment. Initial working LBW estimates for each HS category in each length-segment were generated from the Janmahasatian formula [18]. The model was coded into an electronic worksheet (Microsoft Excel for Mac, 2020) and then underwent the first stage of evaluation.

### Datasets

The National Health and Nutrition Examination Survey (NHANES) datasets from the 1999-2000 to the 2017-2018 surveys were downloaded from the CDC website [19]. The downloaded data included the following variables: sequence number, sex, race, age, TBW, height, body mass index (BMI), mid-arm circumference (MAC), fat mass (FM) and fat free mass (FFM). Data from children under the age of 16 years was excluded, as was data from individuals with incomplete or missing data. The datasets were pooled as follows: the initial model testing was done in the NHANES 2011-2014 surveys, the model calibration was done in the NHANES 1999-2014 surveys, and the model validation was done in the 2015-2018 survey datasets.

### Initial model testing

The initial model was used to generate estimates of IBW and LBW from height and MAC for each individual in the dataset. The data was analysed separately for each length-segment of the tape. Estimates of IBW by the tape were compared against a “standard” IBW calculated using the method of Peterson [20]. Estimates of LBW by the tape were compared against FFM data in the NHANES dataset (measured by dual energy X-ray absorptiometry (DXA)).

The specific outcomes that were evaluated were: mean percentage error (MPE), which represents the estimation bias; the root mean square percentage error (RMSPE) which quantifies the estimation precision; and the percentage of weight estimations that fall within 10% (P10) and 20% (P20) of the reference weight standard, which denotes overall accuracy.

After the initial testing it was evident that two additional habitus score categories were needed to accommodate weight estimations in adults with severe and morbid obesity. The length-segments and MAC cutpoints were modified accordingly. This new model was then calibrated in the larger pooled dataset.

### Model calibration

IBW and LBW estimates generated in the calibration dataset by the refined model were again examined for each length-segment of the tape. The MAC cutpoint values and predicted weight values on the tape were adjusted until a target P20 >95% and P10 >70% (or as close to this as possible) was obtained for each habitus score category in each length-segment. The final model, shown in Table 1, was then subjected to formal validation.

**Table 1.** Final model for the male (upper panel) and female (lower panel) PAWPER XL-MAC adult tape. HS = habitus score, LBW = lean body weight, MAC = mid-arm circumference.

| MALES | HS1 |  | HS2 |  | HS3 |  | HS4 |  | HS5 |  | HS6 |  | HS7 |  | HS8 |  | HS9 |  |
| --- | --- | --- | --- | --- | --- | --- | --- | --- | --- | --- | --- | --- | --- | --- | --- | --- | --- | --- |
| Length | LBW | MAC range | LBW | MAC range | LBW | MAC range | LBW | MAC range | LBW | MAC range | LBW | MAC range | LBW | MAC range | LBW | MAC range | LBW | MAC range |
| (cm) | (kg) | (mm) | (kg) | (mm) | (kg) | (mm) | (kg) | (mm) | (kg) | (mm) | (kg) | (mm) | (kg) | (mm) | (kg) | (mm) | (kg) | (mm) |
| ≤153.0 | **32** | ≤232 | **34** | 233 to 258 | **43** | 259 to 292 | **44** | 293 to 330 | **50** | 331 to 347 | **51** | 348 to 354 | **53** | 355 to 364 | **55** | 365 to 384 | **68** | ≥385 |
| 153.1 to 158.0 | **34** | ≤236 | **37** | 237 to 257 | **42** | 258 to 292 | **49** | 293 to 343 | **55** | 344 to 366 | **59** | 367 to 396 | **60** | 397 to 410 | **61** | 411 to 420 | **63** | ≥421 |
| 158.1 to 162.0 | **37** | ≤228 | **41** | 229 to 269 | **46** | 270 to 297 | **49** | 298 to 327 | **55** | 328 to 370 | **61** | 371 to 392 | **67** | 393 to 417 | **68** | 418 to 429 | **70** | ≥430 |
| 162.1 to 165.0 | **37** | ≤234 | **42** | 235 to 268 | **47** | 269 to 300 | **53** | 301 to 344 | **59** | 345 to 375 | **61** | 376 to 390 | **68** | 391 to 433 | **71** | 434 to 470 | **93** | ≥471 |
| 165.1 to 168.0 | **40** | ≤246 | **45** | 247 to 276 | **50** | 277 to 307 | **55** | 308 to 352 | **62** | 353 to 379 | **66** | 380 to 392 | **67** | 393 to 419 | **72** | 420 to 466 | **80** | ≥467 |
| 168.1 to 171.0 | **41** | ≤236 | **46** | 237 to 277 | **51** | 278 to 304 | **56** | 305 to 347 | **61** | 348 to 379 | **67** | 380 to 405 | **74** | 406 to 434 | **78** | 435 to 480 | **95** | ≥481 |
| 171.1 to 174.0 | **43** | ≤243 | **48** | 244 to 280 | **53** | 281 to 312 | **58** | 313 to 345 | **63** | 346 to 382 | **70** | 383 to 416 | **80** | 417 to 446 | **84** | 447 to 481 | **93** | ≥482 |
| 174.1 to 177.0 | **45** | ≤248 | **50** | 249 to 281 | **55** | 282 to 315 | **60** | 316 to 355 | **67** | 356 to 382 | **72** | 383 to 411 | **77** | 412 to 437 | **82** | 438 to 464 | **91** | ≥465 |
| 177.1 to 180.0 | **47** | ≤255 | **52** | 256 to 286 | **58** | 287 to 324 | **64** | 325 to 360 | **70** | 361 to 400 | **77** | 401 to 419 | **82** | 420 to 478 | **90** | 479 to 497 | **94** | ≥498 |
| 180.1 to 183.0 | **50** | ≤256 | **54** | 257 to 285 | **59** | 286 to 324 | **65** | 325 to 364 | **73** | 365 to 392 | **78** | 393 to 431 | **88** | 432 to 477 | **97** | 478 to 525 | **109** | ≥526 |
| 183.1 to 186.0 | **49** | ≤251 | **54** | 252 to 282 | **60** | 283 to 324 | **67** | 325 to 362 | **74** | 363 to 411 | **84** | 412 to 438 | **91** | 439 to 461 | **96** | 462 to 489 | **108** | ≥490 |
| 186.1 to 189.0 | **52** | ≤263 | **59** | 264 to 294 | **65** | 295 to 333 | **71** | 334 to 369 | **76** | 370 to 409 | **86** | 410 to 442 | **93** | 443 to 469 | **101** | 470 to 502 | **130** | ≥503 |
| 189.1 to 192.0 | **56** | ≤267 | **65** | 268 to 288 | **65** | 289 to 330 | **73** | 331 to 366 | **79** | 367 to 416 | **91** | 417 to 434 | **95** | 435 to 456 | **100** | 457 to 485 | **104** | ≥486 |
| ≥192.1 | **55** | ≤259 | **61** | 260 to 293 | **70** | 294 to 338 | **78** | 339 to 382 | **85** | 383 to 416 | **90** | 417 to 431 | **94** | 432 to 447 | **105** | 448 to 489 | **123** | ≥490 |

| FEMALES | HS1 |  | HS2 |  | HS3 |  | HS4 |  | HS5 |  | HS6 |  | HS7 |  | HS8 |  | HS9 |  |
| --- | --- | --- | --- | --- | --- | --- | --- | --- | --- | --- | --- | --- | --- | --- | --- | --- | --- | --- |
| Length | LBW | MAC range | LBW | MAC range | LBW | MAC range | LBW | MAC range | LBW | MAC range | LBW | MAC range | LBW | MAC range | LBW | MAC range | LBW | MAC range |
| (cm) | (kg) | (mm) | (kg) | (mm) | (kg) | (mm) | (kg) | (mm) | (kg) | (mm) | (kg) | (mm) | (kg) | (mm) | (kg) | (mm) | (kg) | (mm) |
| ≤144.0 | **24** | ≤196 | **26** | 197 to 238 | **29** | 239 to 272 | **33** | 273 to 301 | **35** | 302 to 334 | **37** | 335 to 370 | **43** | 371 to 394 | **44** | 395 to 472 | **46** | ≥473 |
| 144.1 to 147.0 | **24** | ≤217 | **27** | 218 to 244 | **30** | 245 to 270 | **33** | 271 to 305 | **37** | 306 to 342 | **40** | 343 to 375 | **45** | 376 to 394 | **47** | 395 to 436 | **50** | ≥437 |
| 147.1 to 150.0 | **28** | ≤222 | **30** | 223 to 244 | **33** | 245 to 284 | **37** | 285 to 324 | **40** | 325 to 360 | **44** | 361 to 384 | **46** | 385 to 401 | **50** | 402 to 442 | **56** | ≥443 |
| 150.1 to 153.0 | **30** | ≤215 | **32** | 216 to 255 | **34** | 256 to 287 | **37** | 288 to 312 | **40** | 313 to 349 | **45** | 350 to 395 | **48** | 396 to 419 | **55** | 420 to 472 | **58** | ≥473 |
| 153.1 to 156.0 | **30** | ≤223 | **33** | 224 to 253 | **35** | 254 to 291 | **38** | 292 to 321 | **42** | 322 to 345 | **45** | 346 to 405 | **50** | 406 to 427 | **57** | 428 to 473 | **63** | ≥474 |
| 156.1 to 159.0 | **31** | ≤219 | **34** | 220 to 260 | **37** | 261 to 297 | **41** | 298 to 330 | **44** | 331 to 366 | **49** | 367 to 411 | **54** | 412 to 433 | **58** | 434 to 506 | **72** | ≥507 |
| 159.1 to 162.0 | **33** | ≤225 | **36** | 226 to 258 | **39** | 259 to 294 | **42** | 295 to 334 | **47** | 335 to 380 | **53** | 381 to 418 | **59** | 419 to 439 | **60** | 440 to 479 | **68** | ≥480 |
| 162.1 to 165.0 | **34** | ≤226 | **37** | 227 to 262 | **40** | 263 to 294 | **44** | 295 to 334 | **49** | 335 to 380 | **54** | 381 to 418 | **59** | 419 to 450 | **63** | 451 to 483 | **70** | ≥484 |
| 165.1 to 168.0 | **34** | ≤229 | **39** | 230 to 260 | **42** | 261 to 296 | **45** | 297 to 335 | **51** | 336 to 377 | **56** | 378 to 415 | **63** | 416 to 463 | **66** | 464 to 509 | **78** | ≥510 |
| 168.1 to 171.0 | **39** | ≤229 | **41** | 230 to 270 | **44** | 271 to 296 | **47** | 297 to 339 | **52** | 340 to 382 | **58** | 383 to 421 | **61** | 422 to 457 | **68** | 458 to 511 | **82** | ≥512 |
| 171.1 to 174.0 | **39** | ≤237 | **43** | 238 to 269 | **45** | 270 to 302 | **49** | 303 to 337 | **54** | 338 to 374 | **60** | 375 to 425 | **64** | 426 to 461 | **72** | 462 to 497 | **80** | ≥498 |
| 174.1 to 177.0 | **39** | ≤235 | **44** | 236 to 271 | **47** | 272 to 303 | **50** | 304 to 349 | **55** | 350 to 379 | **62** | 380 to 409 | **67** | 410 to 462 | **74** | 463 to 479 | **78** | ≥480 |
| 177.1 to 180.0 | **43** | ≤236 | **46** | 237 to 270 | **49** | 271 to 322 | **54** | 323 to 349 | **57** | 350 to 368 | **60** | 369 to 418 | **68** | 419 to 450 | **73** | 451 to 480 | **78** | ≥481 |
| ≥180.1 | **47** | ≤256 | **48** | 257 to 277 | **49** | 278 to 312 | **55** | 313 to 344 | **63** | 345 to 378 | **69** | 379 to 406 | **69** | 407 to 434 | **77** | 435 to 479 | **85** | ≥480 |

### Model validation

The final model was validated by generating estimated weights in the pooled, unused 2015-2018 NHANES survey datasets. The performance of the model was evaluated in the same way as in the calibration stage. Subgroup analyses by sex, age, race (as defined in the NHANES datasets) and weight-status (determined by the CDC definitions of BMI as underweight, normal weight, overweight, obese, severely obese, and morbidly obese) were also performed.

To be able to compare the PAWPER XL-MAC for adults system with other, previously described, LBW estimation methods, the root mean square error (RMSE) was calculated in the calibration and validation datasets. This is not an ideal statistic to evaluate predictive performance, but it is the only useful outcome measure that has been reported in the few previously published studies.

All data was analysed using Stata Statistical Software (StataCorp. 2019. Stata Statistical Software: Release 16. College Station, TX: StataCorp LLC). A significance level of 0.05 was used throughout.

### Outcome measures

The primary outcome measure was the performance of the model when compared to calculated IBW and measured FFM. A P20 >95% and a P10 >70% was considered to be an acceptable accuracy of estimation, as has previously been suggested for studies on TBW estimation [21, 22].

### Ethics

No ethical approval was required, as this study as no patient participants were included. The data was obtained from the CDC’s online open access databases, which are completely anonymised.

## RESULTS

### Characteristics of study participants

The demographic details and characteristics of the participants included in the calibration and validation studies are shown in Table 2. There were 33215 adults included in the initial calibration study and 5926 in the validation study. There were some important differences between the older derivation datasets and the most recent pooled validation datasets. The validation dataset had a significantly lower proportion of normal weight individuals and a higher proportion of obese, severely obese, and morbidly obese individuals (Chi-squared test, p<0.0001). There was also a significant difference in the ethnic distribution, with a higher proportion of Mexican Americans and lower proportion of Non-Hispanic Whites in the validation dataset (Chi-squared test, p<0.0001).

**Table 2.** Demographic characteristics of the calibration (developmental) and validation datasets. UQ = upper quartile, LQ = lower quartile, BMI = body mass index. The NHANES datasets are not fully representative of the US population, as some subgroups of age and race or ethnicity are oversampled. This dataset therefore contains a higher proportion of Non-Hispanic Black participants, Hispanic participants and Asian participants than the general population. However, the overall age distribution and BMI distribution are very similar to that of the general US population.

|  | CALIBRATION DATASET |  |  | VALIDATION DATASET |  |  |
| --- | --- | --- | --- | --- | --- | --- |
|  | All | Male | Female | All | Male | Female |
| **N (%)** | 33215 | 16857 | 16358 | 5926 | 2886 | 3040 |
| **Age (years)** |  |  |  |  |  |  |
| **Median (UQ, LQ)** | 38 (23, 53) | 37 (22, 53) | 40 (24, 53) | 36 (24, 48) | 35 (24, 47) | 36 (25, 48) |
| **<20 years (%)** | 6043 (18.2) | 3298 (19.6) | 2745 (16.8) | 924 (15.6) | 465 (16.1) | 459 (15.1) |
| **20 to 29.9 years (%)** | 5615 (16.9) | 2926 (17.4) | 2689 (16.4) | 1252 (21.1) | 630 (21.8) | 622 (20.5) |
| **30 to 39.9 years (%)** | 5507 (16.6) | 2797 (16.6) | 2710 (16.6) | 1234 (20.8) | 609 (21.1) | 625 (20.6) |
| **40 to 49.9 years (%)** | 5826 (17.5) | 2814 (16.7) | 3012 (18.4) | 1241 (20.9) | 571 (19.8) | 670 (22.0) |
| **50 to 59.9 years (%)** | 5038 (15.2) | 2463 (14.6) | 2575 (15.7) | 1275 (21.5) | 611 (21.2) | 664 (21.8) |
| **60 to 69.9 years (%)** | 2714 (8.2) | 1324 (7.9) | 1390 (8.5) | 0 | 0 | 0 |
| **70 to 79.9 years (%)** | 1496 (4.5) | 787 (4.7) | 709 (4.3) | 0 | 0 | 0 |
| **≥80 years (%)** | 976 (2.9) | 448 (2.7) | 528 (3.2) | 0 | 0 | 0 |
| **BMI (kgm<sup>-2</sup>)** |  |  |  |  |  |  |
| **Median (UQ, LQ)** | 26.9 (23.2, 31.5) | 26.8 (23.4, 30.5) | 27.1 (23.3, 33.6) | 27.5 (23.4, 32.5) | 27.3 (23.8, 31.5) | 27.8 (23.1, 33.3) |
| **<18.5 kg/m<sup>2</sup> (%)** | 843 (2.5) | 381 (2.3) | 462 (2.8) | 156 (2.6) | 68 (2.4) | 88 (2.9) |
| **18.5 to 24.9 kg/m<sup>2</sup> (%)** | 11369 (34.2) | 5722 (33.9) | 5647 (34.5) | 1882 (31.8) | 897 (31.1) | 985 (32.4) |
| **25 to 29.9 kg/m<sup>2</sup> (%)** | 10451 (31.5) | 6053 (35.9) | 4398 (26.9) | 1750 (29.5) | 964 (33.4) | 786 (25.9) |
| **30 to 34.9 kg/m<sup>2</sup> (%)** | 6008 (18.1) | 2998 (17.8) | 3010 (18.4) | 1147 (19.4) | 565 (19.6) | 582 (19.1) |
| **35 to 39.9 kg/m<sup>2</sup> (%)** | 2685 (8.1) | 1080 (6.4) | 1605 (9.8) | 573 (9.7) | 240 (8.3) | 333 (11.0) |
| **40 to 44.9 kg/m<sup>2</sup> (%)** | 1158 (3.5) | 411 (2.4) | 747 (4.6) | 274 (4.6) | 102 (3.5) | 172 (5.7) |
| **45 to 49.9 kg/m<sup>2</sup> (%)** | 443 (1.3) | 131 (0.8) | 312 (1.9) | 97 (1.6) | 32 (1.1) | 65 (2.1) |
| **≥50 kg/m<sup>2</sup> (%)** | 258 (0.8) | 81 (0.5) | 177 (1.1) | 47 (0.8) | 18 (0.6) | 29 (1.0) |
| **Ethnicity** |  |  |  |  |  |  |
| **Mexican American (%)** | 2049 (15.7) | 1008 (15.6) | 1041 (15.8) | 1074 (18.1) | 508 (17.6) | 566 (18.6) |
| **Other Hispanic (%)** | 1343 (10.3) | 606 (9.4) | 737 (11.2) | 674 (11.4) | 305 (10.6) | 369 (12.1) |
| **Non-Hispanic White (%)** | 4321 (33.1) | 2161 (33.5) | 2160 (32.8) | 1769 (29.9) | 827 (28.7) | 942 (31.0) |
| **Non-Hispanic Black (%)** | 2891 (22.2) | 1399 (21.7) | 1492 (22.6) | 1193 (20.1) | 600 (20.8) | 593 (19.5) |
| **Non-Hispanic Asian (%)** | 1866 (14.3) | 978 (15.1) | 888 (13.5) | 916 (15.5) | 485 (16.8) | 431 (14.2) |
| **Other (%)** | 578 (4.4) | 306 (4.7) | 272 (4.1) | 300 (5.1) | 161 (5.6) | 139 (4.6) |
| **Weight (kg)** |  |  |  |  |  |  |
| **Median (UQ, LQ)** | 76.2 (64.2, 90.4) | 81.1 (70.2, 94.2) | 70.1 (59.3, 84.8) | 76.7 (64.2, 91.4) | 81.4 (70.6, 95.6) | 70.9 (59.2, 86.5) |
| **Height (cm)** |  |  |  |  |  |  |
| **Median (UQ, LQ)** | 167.6 (160.6, 175) | 174.5 (169.3, 179.6) | 161.1 (156.3, 165.9) | 166.7 (159.7, 173.9) | 173.7 (168.6, 178.4) | 160.6 (155.7, 165.3) |

### Validation of IBW estimation by the modified PAWPER XL-MAC method

The PAWPER XL-MAC method, using the HS3 weight to estimate IBW, achieved almost perfect performance in both datasets. In both the calibration and validation datasets the MPE was 2.0%, with a P10 of 99.9% and a P20 of 100%, with no differences across any subgroups.

### Validation of LBW estimation by the modified PAWPER XL-MAC method

The modified PAWPER XL-MAC method exceeded the predefined acceptable outcome criteria in the validation dataset overall, and in every segment-by-segment analysis. The outcomes of the analyses are shown in Table 3. Analysis by sex (male and female) showed a significantly better performance in males in both the calibration and validation datasets (Chi-squared test, p<0.0001). Subgroup analyses by age and race (ethnicity) showed little difference between the groups. In terms of body habitus, the primary accuracy outcome was achieved in all subgroups except extremes of habitus (especially morbidly obese females with a BMI ≥45kg/m^2^). In these participants the accuracy was substantially lower in females than in males (Chi-squared test, p<0.0001). The relationship between MAC and BMI was disrupted in patients with very high BMI, especially in females, which lead to the poorer LBW predictions in these subgroups.

**Table 3.** Results of the weight estimation performance analyses for the calibration (developmental) and validation datasets overall and by subgroups of age, weight status and ethnicity. MPE = mean percentage error, P10 = percentage of weight estimations within 10% of measured fat-free mass, P20 = percentage of weight estimations within 20% of measured fat-free mass.

| LBW | CALIBRATION DATASET (N=33155) |  |  |  |  |  |  |  |  | VALIDATION DATASET (N=5926) |  |  |  |  |  |  |  |  |
| --- | --- | --- | --- | --- | --- | --- | --- | --- | --- | --- | --- | --- | --- | --- | --- | --- | --- | --- |
|  | All |  |  | Male |  |  | Female |  |  | All |  |  | Male |  |  | Female |  |  |
|  | MPE (%) | P10 (%) | P20 (%) | MPE (%) | P10 (%) | P20 (%) | MPE (%) | P10 (%) | P20 (%) | MPE (%) | P10 (%) | P20 (%) | MPE (%) | P10 (%) | P20 (%) | MPE (%) | P10 (%) | P20 (%) |
| ALL | -0.6 | 80.0 | 98.4 | -0.4 | 85.0 | 99.4 | -0.7 | 74.9 | 97.3 | -1.7 | 78.3 | 98.3 | -0.8 | 83.9 | 99.1 | -2.5 | 73.0 | 97.5 |
| AGE |  |  |  |  |  |  |  |  |  |  |  |  |  |  |  |  |  |  |
| <20 years | -1.1 | 81.9 | 98.9 | -0.8 | 85.8 | 99.5 | -1.5 | 77.1 | 98.3 | -0.9 | 82.3 | 98.7 | -0.2 | 88.0 | 98.7 | -1.5 | 76.5 | 98.7 |
| 20 to 29.9 years | -0.8 | 80.6 | 98.5 | 0.1 | 85.4 | 99.1 | -1.7 | 75.3 | 97.7 | -1.3 | 79.6 | 98.2 | 0.0 | 85.9 | 98.7 | -2.6 | 73.3 | 97.7 |
| 30 to 39.9 years | -1.3 | 79.5 | 98.0 | -0.6 | 84.6 | 98.8 | -1.9 | 74.4 | 97.2 | -2.3 | 77.1 | 98.2 | -1.2 | 83.6 | 99.3 | -3.4 | 70.7 | 97.1 |
| 40 to 49.9 years | -1.3 | 79.4 | 98.4 | -0.9 | 84.8 | 99.2 | -1.7 | 74.3 | 97.6 | -2.3 | 76.8 | 98.6 | -1.3 | 80.7 | 99.3 | -3.1 | 73.4 | 98.1 |
| 50 to 59.9 years | -0.6 | 79.3 | 97.7 | -1.0 | 84.5 | 98.6 | -0.2 | 74.3 | 96.9 | -1.7 | 77.1 | 97.8 | -1.5 | 82.5 | 99.3 | -1.8 | 72.1 | 96.4 |
| 60 to 69.9 years | 1.4 | 79.3 | 98.0 | -0.1 | 83.9 | 99.2 | 2.8 | 74.9 | 96.9 | - | - | - | - | - | - | - | - | - |
| 70 to 79.9 years | 2.4 | 77.7 | 96.7 | 1.5 | 83.1 | 98.6 | 3.5 | 71.7 | 94.6 | - | - | - | - | - | - | - | - | - |
| ≥80 years | 2.1 | 76.9 | 96.8 | 2.0 | 79.2 | 97.8 | 2.1 | 75.0 | 96.0 | - | - | - | - | - | - | - | - | - |
| BMI |  |  |  |  |  |  |  |  |  |  |  |  |  |  |  |  |  |  |
| <18.5 kg/m<sup>2</sup> (%) | 5.6 | 73.9 | 96.9 | 5.4 | 79.0 | 97.9 | 5.7 | 69.7 | 96.1 | 6.6 | 69.2 | 94.9 | 6.4 | 76.5 | 95.6 | 6.9 | 63.6 | 94.3 |
| 18.5 to 24.9 kg/m<sup>2</sup> (%) | 1.3 | 81.6 | 98.1 | 1.1 | 85.5 | 99.0 | 1.5 | 77.6 | 97.2 | 1.0 | 81.7 | 98.3 | 1.4 | 85.8 | 99.0 | 0.7 | 77.9 | 97.7 |
| 25 to 29.9 kg/m<sup>2</sup> (%) | -0.4 | 83.9 | 98.8 | -0.4 | 87.7 | 99.3 | -0.4 | 78.7 | 98.2 | -1.2 | 84.7 | 99.1 | -0.5 | 88.2 | 99.3 | -2.0 | 80.5 | 99.0 |
| 30 to 34.9 kg/m<sup>2</sup> (%) | -2.4 | 78.4 | 98.6 | -2.5 | 82.7 | 99.2 | -2.3 | 74.2 | 98.0 | -4.1 | 75.4 | 99.0 | -3.3 | 80.7 | 99.5 | -4.8 | 70.3 | 98.5 |
| 35 to 39.9 kg/m<sup>2</sup> (%) | -3.9 | 71.9 | 97.9 | -3.1 | 79.0 | 98.6 | -4.4 | 67.2 | 97.4 | -6.0 | 68.6 | 97.6 | -4.5 | 78.3 | 99.2 | -7.0 | 61.6 | 96.4 |
| 40 to 44.9 kg/m<sup>2</sup> (%) | -4.1 | 68.1 | 96.8 | -2.7 | 75.2 | 98.5 | -4.9 | 64.3 | 95.9 | -6.0 | 65.0 | 97.8 | -3.7 | 74.5 | 99.0 | -7.4 | 59.3 | 97.1 |
| 45 to 49.9 kg/m<sup>2</sup> (%) | -6.6 | 65.7 | 91.6 | -6.4 | 70.2 | 96.2 | -6.7 | 63.8 | 89.7 | -9.3 | 59.8 | 92.8 | -8.0 | 62.5 | 100 | -10.0 | 58.5 | 89.2 |
| ≥50 kg/m<sup>2</sup> (%) | -7.7 | 56.6 | 90.7 | -6.4 | 61.7 | 92.6 | -8.4 | 54.2 | 89.8 | -9.0 | 46.8 | 85.1 | -3.3 | 66.7 | 94.4 | -12.5 | 34.5 | 79.3 |
| ETHNICITY |  |  |  |  |  |  |  |  |  |  |  |  |  |  |  |  |  |  |
| Mexican American | -2.5 | 77.5 | 98 | -1.8 | 82.1 | 98.5 | -3.2 | 72.9 | 97.6 | -2.7 | 78.1 | 98.9 | -1.8 | 83.9 | 99.2 | -3.4 | 73 | 98.6 |
| Other Hispanic | -1.9 | 79.6 | 98.4 | -1.2 | 84.2 | 98.8 | -2.5 | 75.8 | 98.1 | -2.3 | 78.9 | 98.7 | -1.7 | 82.6 | 99.7 | -2.9 | 75.9 | 97.8 |
| Non-Hispanic White | -0.9 | 80.2 | 97.6 | -0.5 | 85.1 | 98.2 | -1.4 | 75.4 | 97 | -1.6 | 80.3 | 98.1 | -0.9 | 86.5 | 99.2 | -2.1 | 74.8 | 97.2 |
| Non-Hispanic Black | -3.3 | 75 | 97.1 | -2.2 | 82.9 | 98.1 | -4.4 | 67.6 | 96.2 | -3.6 | 74.4 | 97.5 | -2.5 | 84.2 | 99.3 | -4.7 | 64.6 | 95.6 |
| Non-Hispanic Asian | 2.1 | 79.8 | 98.3 | 2.7 | 82 | 98.6 | 1.5 | 77.4 | 98 | 2.1 | 79 | 98.6 | 2.9 | 80 | 98.4 | 1.1 | 78 | 98.8 |
| Other | -1.7 | 77.2 | 97.6 | -1 | 80.4 | 98.4 | -2.6 | 73.5 | 96.7 | -1.9 | 80.3 | 98.7 | -0.8 | 85.7 | 98.8 | -3.1 | 74.1 | 98.6 |

In the analysis of the performance of LBW estimation, the RMSE in the calibration dataset was 3.35 kg (3.33, 3.39), and 3.53 kg (3.45, 3.61) in the validation dataset.

## DISCUSSION

This study presents the first description of a point-of-care weight estimation system that can provide estimates of IBW and LBW for the purpose of weight-based drug dose calculations in adult patients. The new PAWPER XL-MAC system for adults was able to estimate both IBW and LBW with an acceptable degree of accuracy, sufficient to allow for confidence that doses calculated from these scalars would be as accurate as can be achieved in an acute or critical care setting. A previous study of the new PAWPER XL-MAC system for adults showed that estimates of TBW were accurate, with a P10 of 71.3% and a P20 of 96.0% (unpublished data).

### The accuracy of LBW estimates

More than 95% of the estimates were within 20% of actual FFM and nearly 80% of estimates were within 10% of FFM. The accuracy of LBW estimation was similar across BMI-categories in the population, although less accurate in morbidly obese participants. The system performed consistently well in the subgroup analyses of age, gender, and ethnicity.

The most common methods of estimating LBW in a clinical setting are formulas using anthropometric variables such as TBW, height, abdominal circumference, and skinfold measurements. More complex methods include techniques such as bioelectrical impedance [23]. In practice, however, formulas based on TBW and height – such as the Boer, James, Hume, and Janmahasatian formulas – are the methods most used in sick patients [18, 24-27]. Of these formulas, there is no clear evidence or consensus on which is the best, although the Boer and Janmahasatian currently appear to be the most used [24, 28].

The estimation of LBW has been poorly studied and most previous studies have used statistical methodology that is incomplete, or inappropriate for method comparison studies. It is therefore difficult to evaluate their accuracy and to compare them with data from other studies, including this one. However, accuracy as determined by the RMSE metric from the present study (3.5 kg (3.5, 3.6)) compares favourably to that of the Janmahasatian formula. In its original validation study, the Janmahasatian formula achieved a RMSE of 4.4 kg (3.4, 5.2), and a RMSE of 5.2 kg (4.9, 5.4) in the only subsequent study to evaluate its performance [18, 29].

### The accuracy of IBW estimates

This is also the first published report of a point-of-care device that could provide an easy and rapid estimation of IBW without calculations. The accuracy of IBW estimates was virtually perfect with the use of the PAWPER XL-MAC system for adults. The observed accuracy was not surprising, however, as almost all current methods of calculating IBW are determined by, and therefore strongly associated with, length. There is no method that is widely regarded as a gold-standard method for determining IBW, not least because there is no true biologically cogent definition of IBW [5, 30]. Nonetheless, IBW is widely used as a surrogate for LBW in drug dose calculations [31].

Little has changed in the various contemporary methods of calculating IBW since the original derivation of the Devine formulas [32]. Modifications by Robinson *et al* and Miller *et al* resulted in very similar results to the original Devine formula [33, 34]. These methods essentially determined that an ideal weight for an individual should be based on a BMI of 22 kg/m^2^ at their height, which was considered to be the weight resulting in optimum health [31]. More recent methods have added some finesse and ease to the calculation of IBW [5, 35].

### Relevance and implications for weight-based drug dosing

It has been suggested that weight estimates that deviate more than 10% from actual, measured weight could result in treatment that itself is ineffective in underdose or life threatening in overdose [9]. Current evidence suggests that about 30 to 50% of weight estimation errors result in dose errors that reach the patient. Harm may be detected in as many as 10% of these dosing error incidents [2, 36]. Accurate weight estimation is therefore relevant and important.

Given its accuracy, the PAWPER XL-MAC system for adults would be ideal to use in any clinical environment where TBW and LBW cannot be measured, especially if the treatment is time critical. The ability to obtain a rapid, accurate estimate of LBW would open the way to clinicians being able to prescribe drug doses based on LBW, where this would be most appropriate (e.g., propofol, ketamine, rocuronium, procainamide, digoxin, epinephrine, fluid and blood therapy, electrolytes, opioids, and some antibiotics). This would allow a far greater flexibility in, and access to, weight-based drug dose calculations. Drug doses could be correctly scaled and individualised for each patient. This could result in better clinical management as well as more accurate dosing during clinical research related to any aspect of drug therapy.

However, a system that is too complex could be problematic [37]. Since the newly developed PAWPER XL-MAC tape for adults system was specifically designed for use during emergency and critical care, it requires no calculations and is as simple to use with minimal training. Evidence from paediatric use of the PAWPER XL system has suggested that human factor errors are low and that it is easy to use during emergencies [37-39]. There is no reason to think that this should be different in adults, but this will need to be confirmed in future prospective studies.

### The need for new approaches to drug dosing

With an increased ability to determine LBW easily and rapidly at the bedside, new strategies for drug dose scaling in normal weight, overweight and obese patients need to be developed. Future research will also need to further define the best scalars to use for individual drugs and to explore the biological validity and clinical role of scalars such as IBW, adjusted body weight and normalised lean weight [1].

### Limitations

The principal limitation of this study was that measurement of length and MAC by a healthcare provider might not be as accurate as that performed by an expert in anthropometry: the effects of human factor errors require evaluation in real-world or simulation situations in future studies. Secondly, the development of this system from data from the USA means that it might not be representative of any other population. This system would need to be evaluated in a range of populations to establish its accuracy in each. While the NHANES data is not directly representative of the general US population demographics, the subgroup stratification by age and BMI is very similar to that of the general US population.

## Conclusions

The consistent high level of accuracy of IBW and TBW estimations achieved by the modified PAWPER XL-MAC exceeded the pre-determined outcome measures. Estimates were accurate in adults of both sexes, all heights, all ethnic groups, all ages, and most habitus-types. The PAWPER XL-MAC for adults shows promise for being used to aid in guiding drug dosing for drugs with narrow therapeutic windows in obese patients when calculations of LBW would be difficult or excessively time-consuming. A point-of-care system that can accurately estimate TBW, LBW and IBW under difficult circumstances has the capacity to facilitate increased accuracy and appropriateness to individualised drug dosing. It could potentially reduce patient harm from drug dosing errors and could be useful for both clinical practice and research.

## Data Availability

Open access data

## FIGURE LEGENDS

**Supplementary Figure 1** How to use the PAWPER XL-MAC tape. The method of using the tape is the same for children and adults. Panel 1 – the tape is stretched from head to heel using a one-person or, more ideally, a two-person technique. Panel 2 – the segment at which the patient’s heel crosses the line is noted. Panel 3 – mid-arm circumference is measured using the scale provided on the tape itself. Panel 4 – this measurement can be applied in the length segment determined in Step 2. The weight can then be read directly off the tape. This tape only shows total body weight, but the adult version of the tape will have total body weight, ideal body weight and lean body weight data.

**Figure 1** The process used in the development, calibration, and validation of the new PAWPER XL-MAC method for estimating ideal body weight and lean body weight in adults. Although lean body weight (LBW) and fat-free mass (FFM) are often used interchangeably, there are subtle differences. These differences are not important for dose calculations. In this paper LBW is used when referring to a predicted weight, while FFM is used when referring to a DXA-measured weight.

